# Multi-domain AD risk burden and plasma biomarkers in cognitively unimpaired adults

**DOI:** 10.64898/2026.06.11.26355499

**Authors:** Michelle Y Li, Paulina Tolosa-Tort, Margo B Heston, Philip S Insel, Shea J Andrews, the PREVENT-AD research group

## Abstract

**Introduction:** Alzheimer’s disease (AD) pathology accumulates decades before symptom onset, yet how the cumulative effect of genetic, familial, and modifiable lifestyle risk burden jointly affects plasma biomarker levels and trajectories in cognitively unimpaired older adults remains unknown.

**Methods:** We analyzed data from 261 participants in the PREVENT-AD cohort. A composite risk score integrating *APOE* ε4 status, polygenic score, family history, and modifiable/lifestyle risk was examined against six plasma biomarkers using linear regression and linear mixed-effects models.

**Results:** *APOE* ε4 was the strongest predictor of plasma biomarker levels. Higher composite risk burden was associated with elevated ptau_181_, ptau_217_, ptau_217_/Aβ_42_, and GFAP levels, and lower Aβ_42/40_ levels. A higher risk burden was predictive of accelerated ptau_181_ accumulation.

**Discussion:** Cumulative AD risk burden is broadly associated with plasma biomarker levels and specifically predicts accelerated ptau_181_ accumulation in cognitively unimpaired older adults, supporting structured composite risk profiling as a framework for AD risk stratification.

## 1. Introduction

Alzheimer’s disease (AD) pathology involves the accumulation of amyloid-β (Aβ) plaques and neurofibrillary tau tangles that begins decades before cognitive symptom onset [1,2]. This prolonged preclinical phase represents an important window for prevention strategies to take place, including lifestyle modifications and pharmacologic interventions [3]. However, effective preclinical risk stratification requires identifying and classifying individuals according to their likelihood of developing AD prior to symptom onset and remains a significant challenge.

Plasma biomarkers detect early AD-related pathological changes and offer a less invasive and more accessible alternative to cerebrospinal fluid (CSF) or neuroimaging biomarkers [1,2]. Aβ_42/40_ and phosphorylated tau species, specifically ptau_181_ and ptau_217_, are predictive of amyloid PET positivity, and ptau_217_/Aβ_42_ offers higher sensitivity and specificity than either measure alone [4–7]. Glial fibrillary acidic protein (GFAP) and neurofilament light chain (NfL) reflect astrocytic activation and neurodegeneration, respectively, and independently predict longitudinal cognitive decline [8]. However, there is substantial heterogeneity in biomarker levels across cognitively unimpaired (CU) individuals and remains not fully understood.

Genetic, clinical, and lifestyle risk factors contribute to biomarker heterogeneity. Apolipoprotein E (*APOE*) ε4 is the strongest AD genetic risk factor and is associated with accelerated amyloid and tau pathology accumulation [9–11]. Polygenic risk scores (PRS) derived from genome-wide association studies capture the cumulative effect of common genetic variants beyond *APOE* [12]. Family history of AD, especially in first-degree relatives, is also associated with increased risk, independent of genetic factors [13]. Modifiable clinical and lifestyle risk factors such as hypertension, obesity, depression, cognitive activity, physical inactivity, and smoking also account for a large portion of attributable AD risk and can be aggregated into validated polyexposure risk scores (PXS) such as the Cognitive Health and Dementia Risk Assessment (CogDrisk) and the Lifestyle for Brain Health Index (LIBRA2) [14,15].

A cumulative risk burden profile may better capture the reality that AD risk is rarely due to a single cause. Composite scoring methods may improve AD prognostic sensitivity and allow examination of whether these risk components jointly predict biomarker variation, and whether their aggregate effect exceeds what any single domain captures independently [16,17]. We have previously shown that a higher composite burden of *APOE*, PRS, family history, and PXS is associated with increased risk of incident dementia [16,18,19]. However, whether this cumulative burden is also associated with early plasma biomarker trajectories in CU individuals remains unclear.

Here, we address this gap using longitudinal data from the Presymptomatic Evaluation of Experimental or Novel Treatments for Alzheimer’s Disease (PREVENT-AD) to examine 1) cross-sectional associations between a composite AD risk burden model and six plasma biomarkers, and 2) longitudinal biomarker trajectories by risk burden level to test whether cumulative risk predicts biomarker accumulation over time. We hypothesized that higher cumulative risk burden would be associated with elevated plasma biomarkers and accelerated biomarker trajectories over time.

## 2. Methods

### 2.1 Study population and inclusion criteria

This study utilizes data from PREVENT-AD, a longitudinal cohort study conducted at the McGill University Research Centre for Studies in Aging, recruited from the greater Montreal area [20]. Eligible participants have at least one first-degree relative with sporadic AD and were CU at screening, confirmed by a Montreal Cognitive Assessment, Clinical Dementia Rating, and Repeatable Battery for the Assessment of Neuropsychological Status (RBANS) assessments. Annual study visits include neuropsychological testing and blood collection. Clinical progression to mild cognitive impairment (MCI) status was determined through a multidisciplinary consensus review, where neuropsychologists reviewed longitudinal cognitive data for participants with cognitive complaints or participants who scored below 1.5 SD on any RBANS domain or 2 subtests of the Rey Auditory Verbal Learning Test, blinded to genetics, imaging and biomarker information. Suspected cases were then discussed and confirmed with a cognitive neurologist and neuropsychiatrist. Dementia status was determined through clinical diagnosis or caregiver report. Aβ PET was available for 232 participants out of the 348 PREVENT-AD participants; a cohort-specific PET centiloid positivity threshold was determined to be 22.32 as previously described [21].

For this analysis, participants were required to have at least two plasma biomarker observations and genetic data including *APOE* and genome-wide genotyping. Of the 348 PREVENT-AD participants, 261 met inclusion criteria and formed the analytic sample. The investigators of the PREVENT-AD program contributed to the design and implementation of PREVENT-AD and/or provided data but did not participate in data analysis or writing of this report. Written informed consent was obtained from all participants, and all research procedures were approved by the Institutional Review Board at McGill University.

### 2.2 Plasma biomarkers

Plasma biomarkers of AD pathology were measured from blood samples collected at study visits and analyzed at the Clinical Neurochemistry Laboratory at the University of Gothenburg. Aβ_42/40_, GFAP, and NfL were analyzed using the commercial Neurology 4-plex E kit (503105; Quanterix) across 1,135 different observations across annual visits. Ptau_181_ and ptau_217_ were analyzed using in-house single molecule array assays at 1,106 and 841 observations across annual visits, respectively. The ptau_217_/Aβ_42_ ratio was manually calculated from the ptau_217_ and Aβ_42_ measures. GFAP, NfL, ptau_181_, ptau_217_, and the ptau_217_/Aβ_42_ ratio were log-transformed prior to analyses to address skewness. The Aβ_42/40_ ratio was analyzed on its original scale.

### 2.3 Risk factors

#### 2.3.1 Genetic risk factors

Participants were classified as *APOE* ε4 carriers if they carried at least one ε4 allele and as non-carriers otherwise. Additionally, 74 independent genome-wide significant single nucleotide polymorphisms (SNPs) previously associated with AD/ADRD in participants of European-descent (64,498 cases; 46,828 proxy cases; 677,663 controls) were available [22]. An AD-PRS was computed as a weighted sum of risk alleles across matched loci, using beta coefficients as weights. Participants in the top quartile of the analysis cohort were classified as high polygenic risk and all others as low polygenic risk [23,24].

#### 2.3.2 Family history risk factor

All PREVENT-AD participants had at least one first-degree relative with AD as a screening criterion, thus information on which first-degree relative(s) as well as the number of extended family members with AD was used to generate the family history risk burden indicator. Family history was classified as high risk if the participant reported 2 or more first-degree relatives with a history of AD, reflecting evidence that a greater number of affected relatives corresponds with a higher familial AD risk beyond the PREVENT-AD inclusion criteria [13].

#### 2.3.3 Polyexposure risk scores (PXS)

Two validated clinical risk scores were used to assess modifiable risk burden and were modified to fit the available data (Table S1).

CogDrisk is a dementia risk assessment tool that integrates 16 risk factors into a single score, spanning demographic, clinical, and lifestyle domains [14]. A modified CogDrisk score was computed from PREVENT-AD assessment and demographic data, where available. 14 out of the 16 CogDrisk items were included: age and sex, education, hypertension, high cholesterol, midlife obesity, physical inactivity, diabetes, depression, smoking, cognitive engagement, traumatic brain injury, insomnia, atrial fibrillation, and social engagement. Diet and stroke were not available. Scores were calculated following the original CogDrisk weighting scheme for available items.

LIBRA2 is a 15-item index developed to quantify modifiable risk and protective factors associated with AD [15]. Compared to CogDrisk, LIBRA2 does not incorporate demographic variables (age, sex, or education) in its scoring as it focuses primarily on modifiable factors. 11 out of the 15 LIBRA2 items were included: hypertension, hypercholesterolemia, obesity, regular physical activity, diabetes, depression, smoking, high cognitive activity, sleep disturbances, low social participation, and hearing impairment. High alcohol intake, diet, coronary heart disease, and chronic heart disease were not available. Scores were calculated following the original LIBRA2 weighting scheme for available items.

Participants with incomplete clinical and lifestyle data within the available CogDrisk and LIBRA2 items at more than 25% were excluded (n=25). Random forest-based imputation was then used to impute variables with partial missingness (<25%) using missForest [25]. The out-of-bag error estimates given were a normalized root mean squared error of 0.027 and the proportion of falsely classified variables of 0.210. Imputation quality was further assessed by calculating the standard deviation (SD) ratio of the imputed clinical risk scores against the scores constructed from complete cases. SD ratios indicated acceptable performance (CogDrisk: SD=1.013; LIBRA2: SD=1.007), so imputed values were used in analysis and scoring.

Scores 1 SD above the cohort mean for each clinical risk score were determined to be “high risk” (CogDrisk: ≥8; LIBRA2: ≥7.3).

### 2.4 Composite risk burden score

A composite risk burden score was constructed by summing binary indicators across the four risk domains for each participant: 1) *APOE* ε4 carrier status, 2) PRS, 3) family history burden, and 4) PXS. Each participant received two composite scores, one CogDrisk-based and one LIBRA2-based. The primary analysis used CogDrisk as it incorporates demographic factors. Scores range from 0 to 4 and are treated as a continuous predictor, assuming a linear increase in biomarker level per additional risk indicator. All four components are weighted equally, reflecting an additive burden assumption [16].

### 2.5 Statistical Analysis

Participant characteristics were assessed using Kruskal-Wallis rank sum test for continuous variables and Pearson’s Chi-squared tests for categorical variables across cognitive progression status.

Statistical significance was set at α=0.05 (two-sided). Unadjusted p-values were reported as an exploratory analysis. To account for multiple comparisons across the 6 biomarkers and 2 composite models, p-values were also adjusted using the Hochberg procedure within each analysis type [26]. PXSs were z-scored prior to inclusion in multivariable models, so beta coefficients reflect the change per standard deviation of the risk score. All statistical analyses were conducted in R (v4.4.3). LMMs were estimated using the lme4 package, and P-values for linear mixed-effects models were estimated using Satterthwaite’s approximation for denominator degrees of freedom via the lmerTest R package [27]. Code used for analyses is available on GitHub (github.com/AndrewsLabUCSF/PREVENTAD-biomarker-trajectories).

#### 2.5.1 Cross-sectional analysis

To estimate the independent contribution of each risk component and the composite association of cumulative risk burden with plasma biomarker levels at the last visit, we fit two sets of linear regression models. First, multivariable linear regression models were fit with *APOE*ε4, family history burden, PRS, and PXS as predictors. Second, separate models tested the association between cumulative risk burden and each plasma biomarker, with the composite risk score entered as the primary predictor. Models were fit separately using CogDrisk-based and LIBRA2-based PXS/composite scores. LIBRA2 models were adjusted for age, sex, and years of education because these variables are not included in the LIBRA2 score.

#### 2.5.2 Longitudinal analysis

To estimate the independent contribution of each risk component and the composite association of cumulative risk burden with longitudinal plasma biomarker trajectories, we fit two sets of linear mixed-effects (LME) models. First, multivariable models included *APOE* ε4, family history burden, PRS, PXS, and their corresponding interactions with time as simultaneous predictors. Second, separate models tested the association between cumulative risk burden and each plasma biomarker trajectory, with the composite risk score, time, and their interaction entered as the primary predictors. Time was defined as years from study entry and centered at the cohort mean. Models included participant-level random intercepts and random slopes for time; for ptau_217_/Aβ_42_ models, only random intercepts were included because random slopes were not estimable due to too few ptau_217_ measurements across the cohort. Models were fit separately using CogDrisk-based and LIBRA2-based PXS/composite scores. LIBRA2 models were adjusted for age, sex, and years of education because these variables are not included in the LIBRA2 score.

#### 2.5.3 Sensitivity analyses

Sensitivity analyses excluded participants who progressed to MCI or dementia during follow-up to assess whether findings were driven by clinical progression.

## 3. Results

### 3.1 Demographic characteristics

Of the 348 PREVENT-AD participants, 261 met inclusion and exclusion criteria (Table 1). The mean age at baseline was 62.8 (SD: 5.0), 71% were female, and 99% were of European descent. 77 participants progressed to MCI or dementia status. Compared to participants who remained CU, those who progressed to MCI or dementia were older at baseline, had higher CogDrisk scores, and higher ptau_217_ and ptau_217_/Aβ_42_ levels. *APOE* ε4 carrier status was the most prevalent individual risk factor, and the largest intersecting subgroup in both the CogDrisk and LIBRA2 models comprised of participants with *APOE* ε4 as their sole risk factor (n=55 and n=46, respectively) (Figure 1). Of the 226 participants with amyloid-PET data, 47 were amyloid positive.

**Figure 1.**
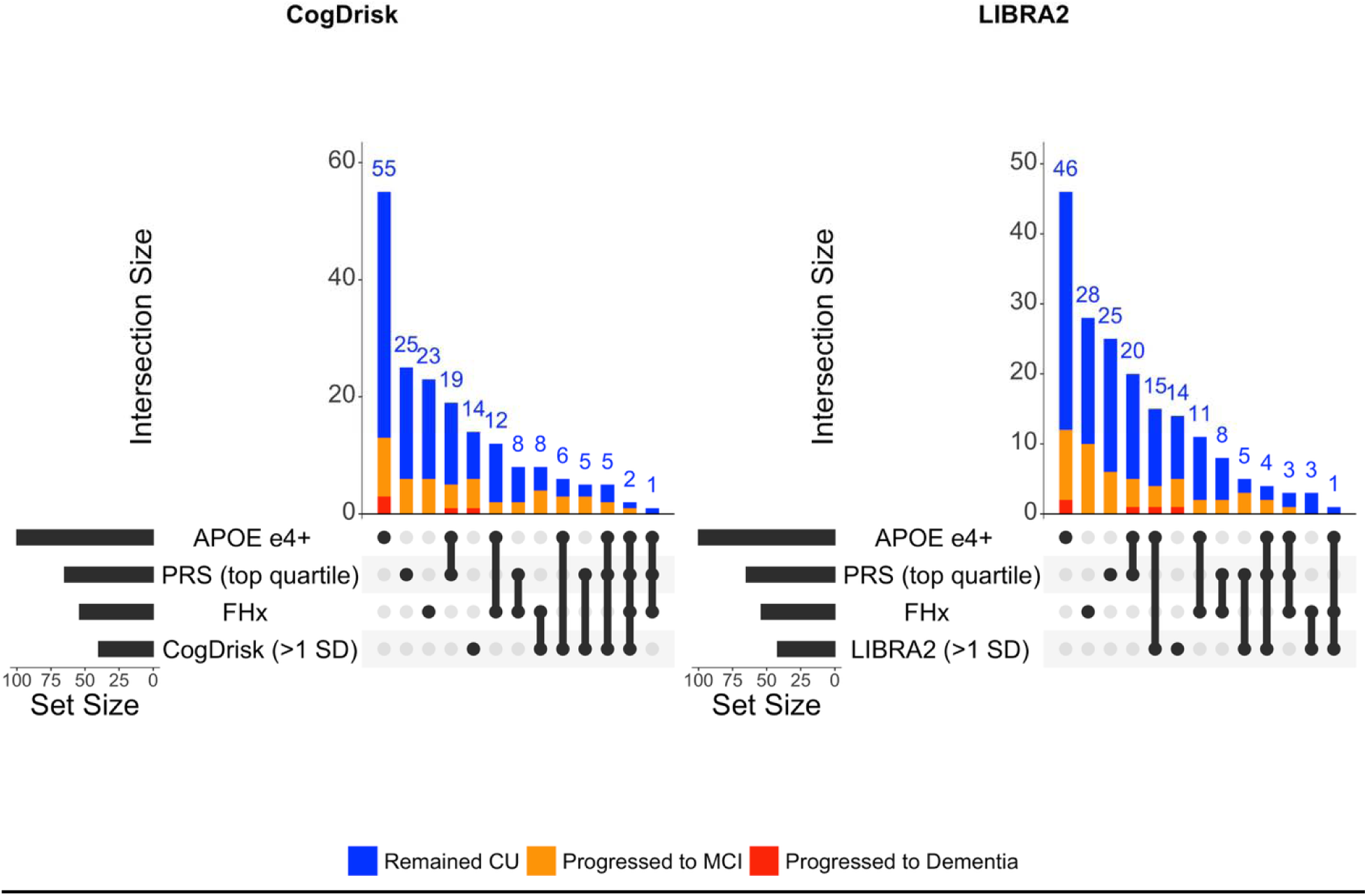
Upset plots showing the distribution and intersections of AD risk indicators at baseline among participants who remained CU and those who progressed to MCI status or dementia. Vertical bars show the number of individuals with the respective combinations of risk indicators. Risk indicators shown are 1) *APOE* ε4 carrier status, 2) high PRS (top quartile), 3) FHx (≥2 first-degree relatives), and 4) high CogDrisk or LIBRA2 score (1 SD above sample mean). Abbreviations: AD, Alzheimer’s disease; *APOE*, apolipoprotein E; CogDrisk, Cognitive Health and Dementia Risk Assessment; CU, cognitively unimpaired; FHx, family history of AD; LIBRA2, Lifestyle for Brain Health Index; MCI, mild cognitive impairment; PREVENT-AD, Presymptomatic Evaluation of Experimental or Novel Treatments for Alzheimer’s Disease; PRS, polygenic risk score; SD, standard deviation.

**Table 1.** Baseline Characteristics of PREVENT-AD Sample. Stratified by Cognitive Progression Status

| Characteristic | Overall<br>N = 261<sup>1</sup> | Remained CU<br>N = 184<sup>1</sup> | Progressed to<br>MCI/dementia<br>N = 77<sup>1</sup> | p-value<sup>2</sup> |
| --- | --- | --- | --- | --- |
| Demographics |  |  |  |  |
| Age (years) | 62.8 (5.0) | 62.0 (4.7) | 64.7 (5.4) | <0.001 |
| Female | 186 (71%) | 132 (72%) | 54 (70%) | >0.9 |
| Years of education | 15 (3) | 15 (3) | 15 (4) | 0.081 |
| Caucasian | 258 (99%) | 182 (99%) | 76 (99%) | >0.9 |
| Amyloid-PET +<sup>3</sup> | 47 (25%) | 25 (19%) | 22 (39%) | 0.007 |
| AD risk indicators |  |  |  |  |
| APOE e4+ | 100 (38%) | 70 (38%) | 30 (39%) | >0.9 |
| PRS ≥75% | 65 (25%) | 45 (24%) | 20 (26%) | >0.9 |
| Family history of AD | 54 (21%) | 39 (21%) | 15 (19%) | 0.9 |
| CogDrisk ≥8 | 40 (15%) | 20 (11%) | 20 (26%) | 0.004 |
| LIBRA2 ≥7.3 | 42 (16%) | 26 (14%) | 16 (21%) | 0.3 |
| AD risk factors |  |  |  |  |
| APOE genotype |  |  |  | 0.8 |
| e3/e3 | 128 (49%) | 89 (48%) | 39 (51%) |  |
| e2+ | 33 (13%) | 25 (14%) | 8 (10%) |  |
| *e4+* | 100 (38%) | 70 (38%) | 30 (39%) |  |
| **FDRs with AD** |  |  |  | **0.8** |
| *1* | 207 (79%) | 145 (79%) | 62 (81%) |  |
| *2* | 48 (18%) | 34 (18%) | 14 (18%) |  |
| *3* | 4 (1.5%) | 3 (1.6%) | 1 (1.3%) |  |
| *4* | 2 (0.8%) | 2 (1.1%) | 0 (0%) |  |
| **PRS** | 4.87 (0.36) | 4.87 (0.37) | 4.85 (0.33) | **0.6** |
| **CogDrisk score** | 2.8 (4.6) | 2.2 (4.1) | 4.1 (5.3) | **0.017** |
| **LIBRA2 score** | 3.3 (4.0) | 2.8 (4.1) | 4.3 (3.6) | **0.006** |
| **Risk indicator burden** |  |  |  |  |
| **CogDrisk** |  |  |  | **0.7** |
| *0* | 78 (30%) | 59 (32%) | 19 (25%) |  |
| *1* | 117 (45%) | 82 (45%) | 35 (45%) |  |
| *2* | 58 (22%) | 38 (21%) | 20 (26%) |  |
| *3* | 6 (2.3%) | 4 (2.2%) | 2 (2.6%) |  |
| *4* | 2 (0.8%) | 1 (0.5%) | 1 (1.3%) |  |
| **LIBRA2** |  |  |  | **0.8** |
| *0* | 78 (30%) | 58 (32%) | 20 (26%) |  |
| *1* | 113 (43%) | 77 (42%) | 36 (47%) |  |
| *2* | 62 (24%) | 44 (24%) | 18 (23%) |  |
| *3* | 8 (3.1%) | 5 (2.7%) | 3 (3.9%) |  |

| Baseline biomarkers |  |  |  |  |
| --- | --- | --- | --- | --- |
| **Aβ42/40 ratio** | 0.069 (0.013) | 0.069 (0.013) | 0.069 (0.013) | 0.5 |
| **GFAP (pg/mL)** | 96 (43) | 92 (39) | 105 (50) | 0.061 |
| **NfL (pg/mL)** | 15.4 (6.4) | 14.9 (6.4) | 16.4 (6.3) | 0.040 |
| **ptau181 (pg/mL)** | 6.85 (5.12) | 6.96 (5.93) | 6.62 (2.45) | 0.3 |
| **ptau217 (pg/mL)** | 2.26 (1.09) | 2.12 (1.05) | 2.59 (1.14) | <0.001 |
| **ptau217/Aβ42 ratio** | 0.37 (0.21) | 0.35 (0.20) | 0.41 (0.24) | 0.059 |

<sup>1</sup>Mean (SD) for continuous; n (%) for categorical
<sup>2</sup>Kruskal-Wallis rank sum test; Pearson's Chi-squared test
<sup>3</sup>Amyloid PET data was available for 226 participants
\* Abbreviations: Aβ, amyloid beta; AD, Alzheimer's disease; APOE, apolipoprotein E; CogDrisk, Cognitive Health and Dementia Risk Assessment; CU, cognitively unimpaired; FDR, first-degree relative; GFAP, glial fibrillary acidic protein; LIBRA2, Lifestyle for Brain Health Index; MCI, mild cognitive impairment; NfL, neurofilament light chain; PRS, polygenic risk score; ptau, phosphorylated tau

### 3.2 Higher AD composite risk burden is associated with higher plasma biomarker levels

We first evaluated the joint association of each risk component with plasma biomarkers at last visit (Figure 2, <u>Table S2</u>). *APOE* ε4 was significantly associated with higher GFAP, ptau_181_, ptau_217_, and ptau_217_/Aβ_42_ levels, and lower Aβ_42/40_ levels. A higher CogDrisk score was associated with higher GFAP, NfL, and ptau_217_ levels (GFAP: β=0.073 [0.014, 0.131], p=0.016; NfL: β=0.069 [0.019, 0.119], p=0.007; ptau_217_: β=0.083 [0.027, 0.140], p=0.004). A higher PRS was associated with higher GFAP levels (β=0.181 [0.016, 0.346], p=0.039). Family history of AD was not associated with any plasma biomarkers. Findings were similar in the LIBRA2-based multivariable linear regression model, except the PRS had significant associations with NfL (β=0.140 [0.011, 0.268], p=0.035). LIBRA2 was not associated with biomarker levels.

**Figure 2.**
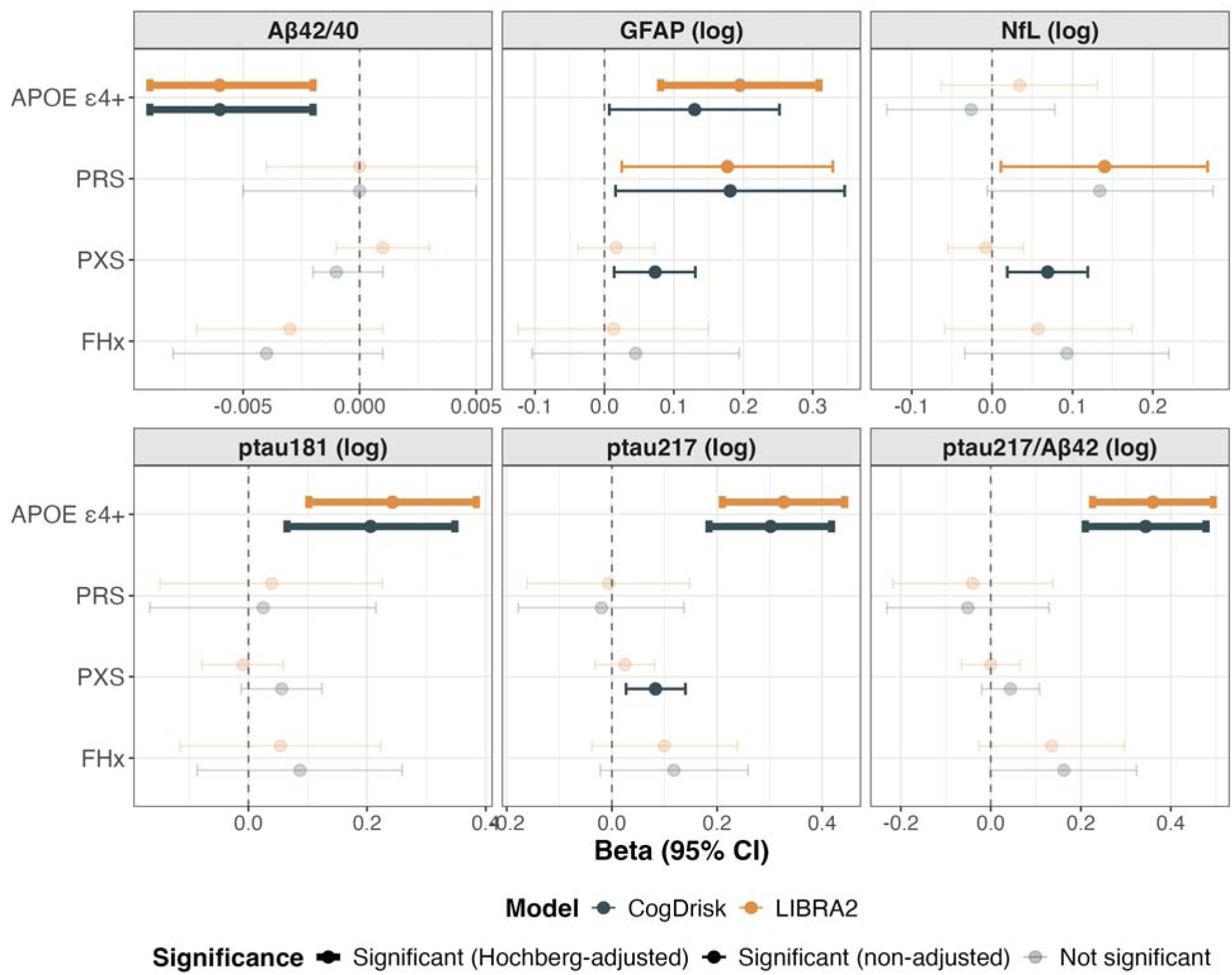
Coefficient plot of multivariate analysis of AD risk burden indicators effects on plasma biomarker levels at last visit among PREVENT-AD participants. Risk indicators: *APOE* ε4 carrier status, PRS, PXS, and family history of AD. Separate models fit for CogDrisk and LIBRA2 as the PXS. LIBRA2 models adjusted for age, sex, and education. Darker points and thicker error bars denote Hochberg-adjusted significance (< 0.05); faded points denote non-significance. Plasma biomarkers: Aβ_42/40_, GFAP, NfL, ptau_181_, ptau_217_, ptau_217_/Aβ_42_. Abbreviations: Aβ, amyloid beta; AD, Alzheimer’s disease; *APOE*, apolipoprotein E; CI, confidence interval; CogDrisk, Cognitive Health and Dementia Risk Assessment; GFAP, glial fibrillary acidic protein; LIBRA2, Lifestyle for Brain Health Index; MCI, mild cognitive impairment; NfL, neurofilament light chain; PRS, polygenic risk score; ptau, phosphorylated tau; PREVENT-AD, Presymptomatic Evaluation of Experimental or Novel Treatments for Alzheimer’s Disease; PXS, polyexposure score.

After Hochberg correction, only associations between *APOE* ε4 and higher ptau_181_, ptau_217_, and ptau_217_/Aβ_42_ levels, and lower Aβ_42/40_ levels, remained significant <u>(Table S2</u>). Additionally, the association between *APOE* ε4 and higher GFAP levels remained significant in a LIBRA2-based model. All other associations lost significance after Hochberg correction. When excluding MCI progressors, *APOE* ε4 carrier status remained associated with higher ptau_217_ and ptau_217_/Aβ_42_ levels, and survived Hochberg correction.

Next, we evaluated the association of composite risk burden with plasma biomarker levels at last visit (Figure 3, <u>Table S3</u>). A higher CogDrisk-based composite risk burden was significantly associated with higher GFAP, NfL, ptau_181_, ptau_217_, and ptau_217_/Aβ_42_ levels, and lower Aβ_42/40_ levels (<u>Table S3</u>). In LIBRA2-based composite models, effect sizes were generally comparable, although the association with NfL was non-significant. All CogDrisk-based associations except NfL remained significant after Hochberg correction. The association between a higher LIBRA2-based composite risk burden and higher GFAP levels and lower Aβ_42/40_ levels lost significance after Hochberg correction. When excluding MCI progressors, higher composite risk burden was not associated with any biomarker level.

**Figure 3.**
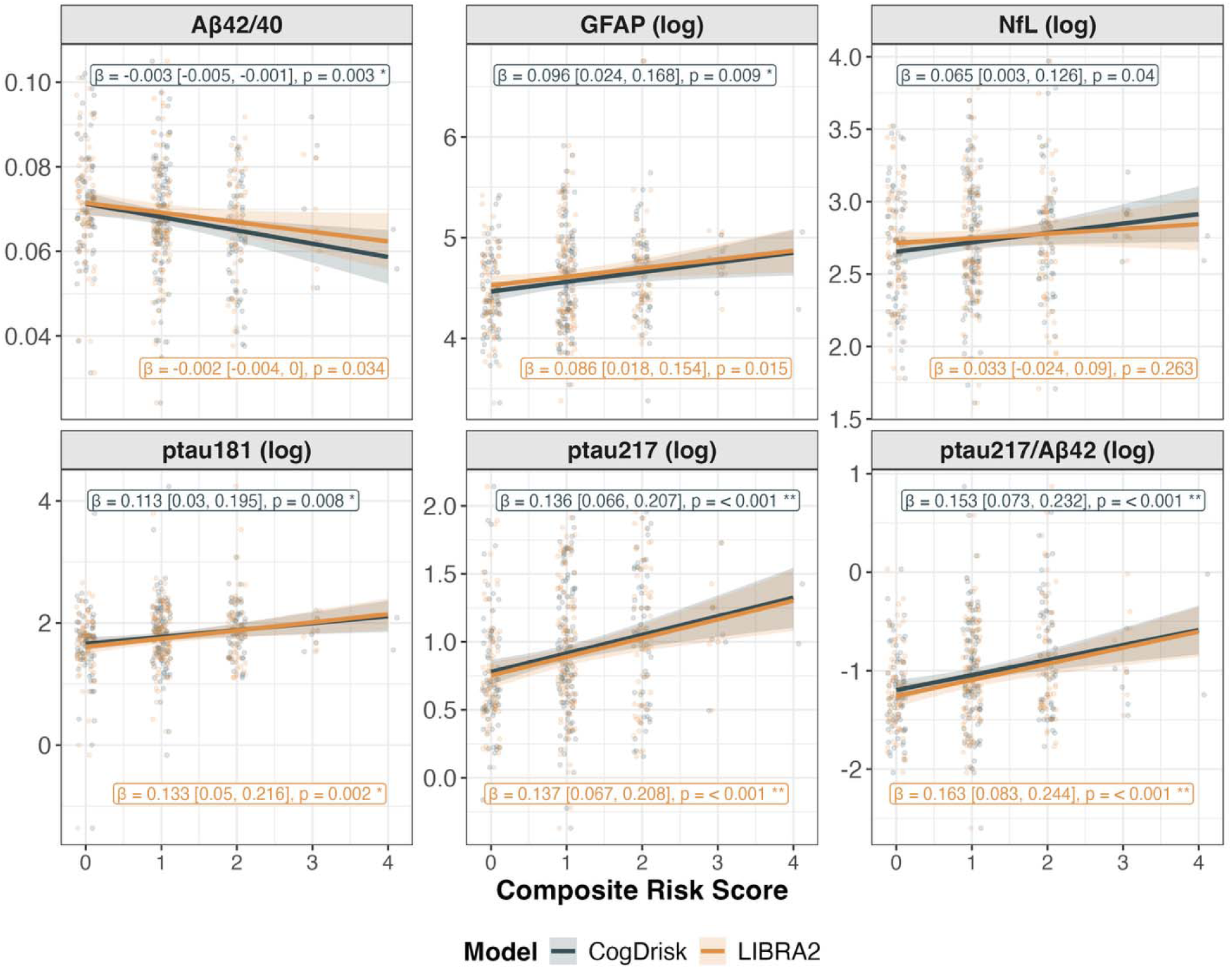
Cross-sectional association between composite AD risk score and plasma biomarker levels among PREVENT-AD participants. Jittered points represent individual observations. Lines show model-predicted values with 95% CIs from linear regression. The composite risk score (0–4) reflects the count of elevated binary risk indicators: *APOE* ε4 carrier status, high PRS (top quartile), family history of AD (≥2 first-degree relatives), and high CogDrisk or LIBRA2 score (1 SD above sample mean). Separate models fit for CogDrisk-Composite and LIBRA2-Composite models. LIBRA2 models adjusted for age, sex, and education. Unadjusted p-values are shown; adjusted p-values can be found in Table S3. Plasma biomarkers: Aβ□□/□□, GFAP, NfL, ptau□□□, ptau□□□, ptau□□□/Aβ□□. Abbreviations: Aβ, amyloid beta; AD, Alzheimer’s disease; *APOE*, apolipoprotein E; β, beta coefficient; CogDrisk, Cognitive Health and Dementia Risk Assessment; CI, confidence interval; GFAP, glial fibrillary acidic protein; LIBRA2, Lifestyle for Brain Health Index; MCI, mild cognitive impairment; NfL, neurofilament light chain; p, p-value; PRS, polygenic risk score; ptau, phosphorylated tau; PREVENT-AD, Presymptomatic Evaluation of Experimental or Novel Treatments for Alzheimer’s Disease; SD, standard deviation.

### 3.3 Higher AD composite risk burden is associated with accelerated ptau_181_ accumulation

We next evaluated the association between AD risk indicators and baseline plasma biomarker levels and trajectories across follow-up using LME models (Figure 4, <u>Table S4</u>). In multivariable LME models including individual risk indicators, *APOE* ε4 was significantly associated with higher GFAP, ptau_181_, ptau_217_, and ptau_217_/Aβ_42_ levels, and lower Aβ_42/40_ levels (<u>Table S5</u>). A higher PRS was associated with higher GFAP and NfL levels. A higher CogDrisk score was associated with higher GFAP (β=0.078 [0.021, 0.134], p=0.008), NfL (β=0.078 [0.030, 0.126], p=0.002), and ptau_217_ levels (β=0.073 [0.017, 0.129], p=0.012). After Hochberg correction, *APOE* ε4 maintained associations but PRS associations with GFAP and NfL lost significance. Additionally, CogDrisk score maintained associations with NfL levels, but its associations with GFAP and ptau_217_ lost significance. After excluding MCI progressors, *APOE* ε4 remained associated with higher ptau_217_/Aβ_42_ levels and survived Hochberg correction. *APOE* ε4 also remained associated with higher GFAP and ptau_217_ levels in LIBRA2-based models, and survived Hochberg correction.

**Figure 4.**
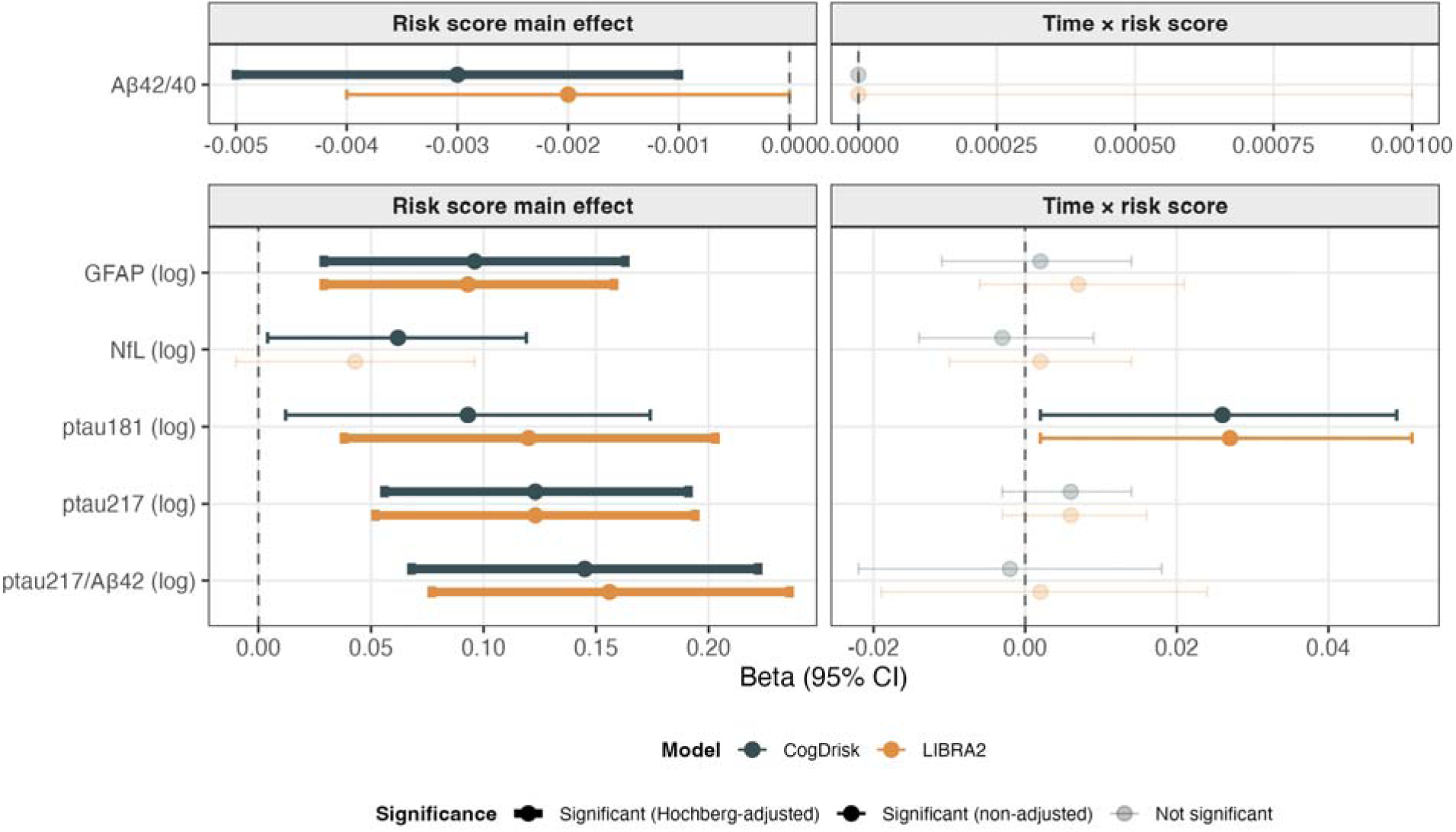
Coefficient plot of linear mixed-effects models examining associations between AD risk burden indicators and plasma biomarker levels across longitudinal follow-up among PREVENT-AD participants. The composite risk score (0–4) reflects the count of elevated binary risk indicators: *APOE* ε4 carrier status, high PRS (top quartile), family history of AD (≥2 first-degree relatives), and high CogDrisk or LIBRA2 score (1 SD above sample mean). Each model included a risk score × time interaction term and was fit with random intercepts and slopes per participant. Left panel shows the conditional main effect of risk score on biomarker levels. Right panel shows the risk score × time interaction coefficient. Aβ□□/□□ shown separately on its own scale. Separate models fit for CogDrisk-Composite and LIBRA2-Composite models. LIBRA2-Composite models adjusted for age, sex, and education. Darker points and thicker error bars denote Hochberg-adjusted significance (< 0.05); faded points denote non-significance. Plasma biomarkers: Aβ□□/□□, GFAP, NfL, ptau□□□, ptau□□□, ptau□□□/Aβ□□. Abbreviations: Aβ, amyloid beta; AD, Alzheimer’s disease; *APOE*, apolipoprotein E; CogDrisk, Cognitive Health and Dementia Risk Assessment; CI, confidence interval; GFAP, glial fibrillary acidic protein; LIBRA2, Lifestyle for Brain Health Index; NfL, neurofilament light chain; PRS, polygenic risk score; ptau, phosphorylated tau; PREVENT-AD, Presymptomatic Evaluation of Experimental or Novel Treatments for Alzheimer’s Disease; SD, standard deviation.

For biomarker trajectories, higher PRS was associated with accelerated NfL accumulation (β=0.031 [0.006, 0.056], p=0.018) and *APOE* ε4 carrier status with accelerated ptau_181_ accumulation (β=0.057 [0.017, 0.097], p=0.006). Findings were similar in the LIBRA2-based multivariable LME model, except the PRS also had significant associations with reduction in Aβ_42/40_ levels (β=0.001 [0.000, 0.002], p=0.039). Additionally, a higher LIBRA2 score was associated with accelerated NfL accumulation (β=0.011 [0.002, 0.020], p=0.023). After Hochberg correction, only *APOE* ε4 maintained association with ptau_181_ levels in a LIBRA2-based multivariable LME model. When excluding MCI progressors, no risk components were associated with rate of change in any biomarker level.

In CogDrisk-composite models, higher cumulative risk burden was significantly associated with higher GFAP, NfL, ptau_217_, and ptau_217_/Aβ_42_ levels, and lower Aβ_42/40_ levels at baseline (Figure 4, <u>Table S4</u>). Higher composite risk burden predicted an accelerated accumulation of ptau_181_ (CogDrisk: β=0.026, p=0.033; LIBRA2: β=0.027, p=0.033) (Figure 5, Figure 6). Effect sizes were comparable across both CogDrisk-based and LIBRA2-based composite models. After Hochberg correction, associations between higher composite risk burden and higher GFAP levels remained significant. Associations between higher LIBRA2-based risk burden and lower Aβ_42/40_ levels lost significance. Associations between higher CogDrisk-based risk burden and higher NfL and ptau_181_ levels also lost significance. No significant associations were found between risk burden and biomarker levels after excluding MCI progressors.

**Figure 5.**
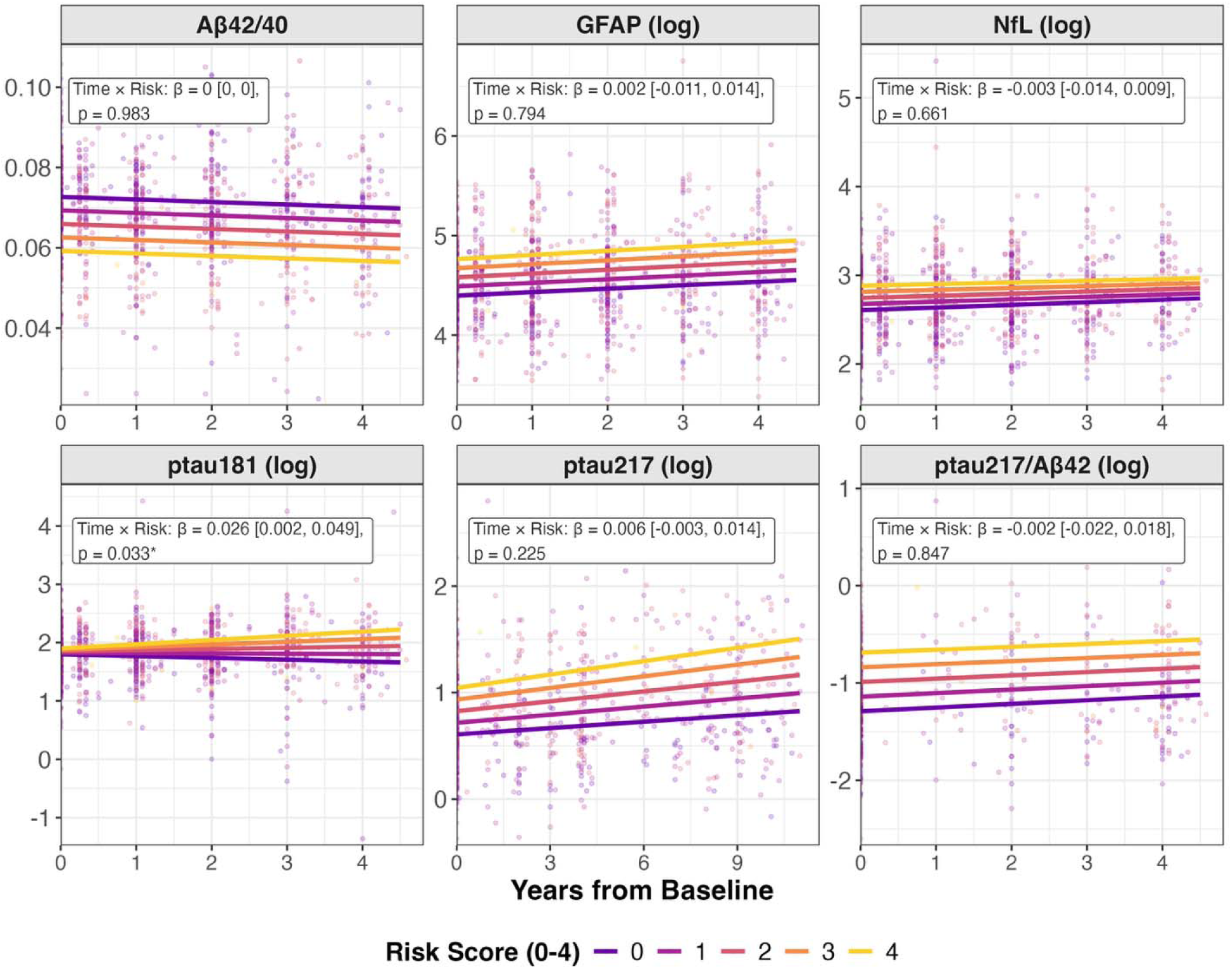
Predicted plasma biomarker trajectories by CogDrisk-based composite AD risk score among PREVENT-AD participants. Points represent individual observations colored by risk score. Lines show population-average predicted trajectories from linear mixed-effects models (biomarker ∼ years × composite risk score + (1 + years | participant)). Unadjusted p-values are shown; adjusted p-values can be found in Table S4. The composite risk score (0–4) reflects the count of elevated binary risk indicators: *APOE* ε4 carrier status, high PRS (top quartile), family history of AD (≥2 first-degree relatives), and high CogDrisk score (1 SD above sample mean). Predictions are generated at a reference demographic profile (female sex, mean baseline age, mean years of education). Plasma biomarkers: Aβ_42/40_, GFAP, NfL, ptau_181_, ptau_217_, ptau_217_/Aβ_42_. Abbreviations: Aβ, amyloid beta; AD, Alzheimer’s disease; *APOE*, apolipoprotein E; β, beta coefficient; CogDrisk, Cognitive Health and Dementia Risk Assessment; GFAP, glial fibrillary acidic protein; NfL, neurofilament light chain; p, p-value; PRS, polygenic risk score; ptau, phosphorylated tau; PREVENT-AD, Presymptomatic Evaluation of Experimental or Novel Treatments for Alzheimer’s Disease; SD, standard deviation.

**Figure 6.**
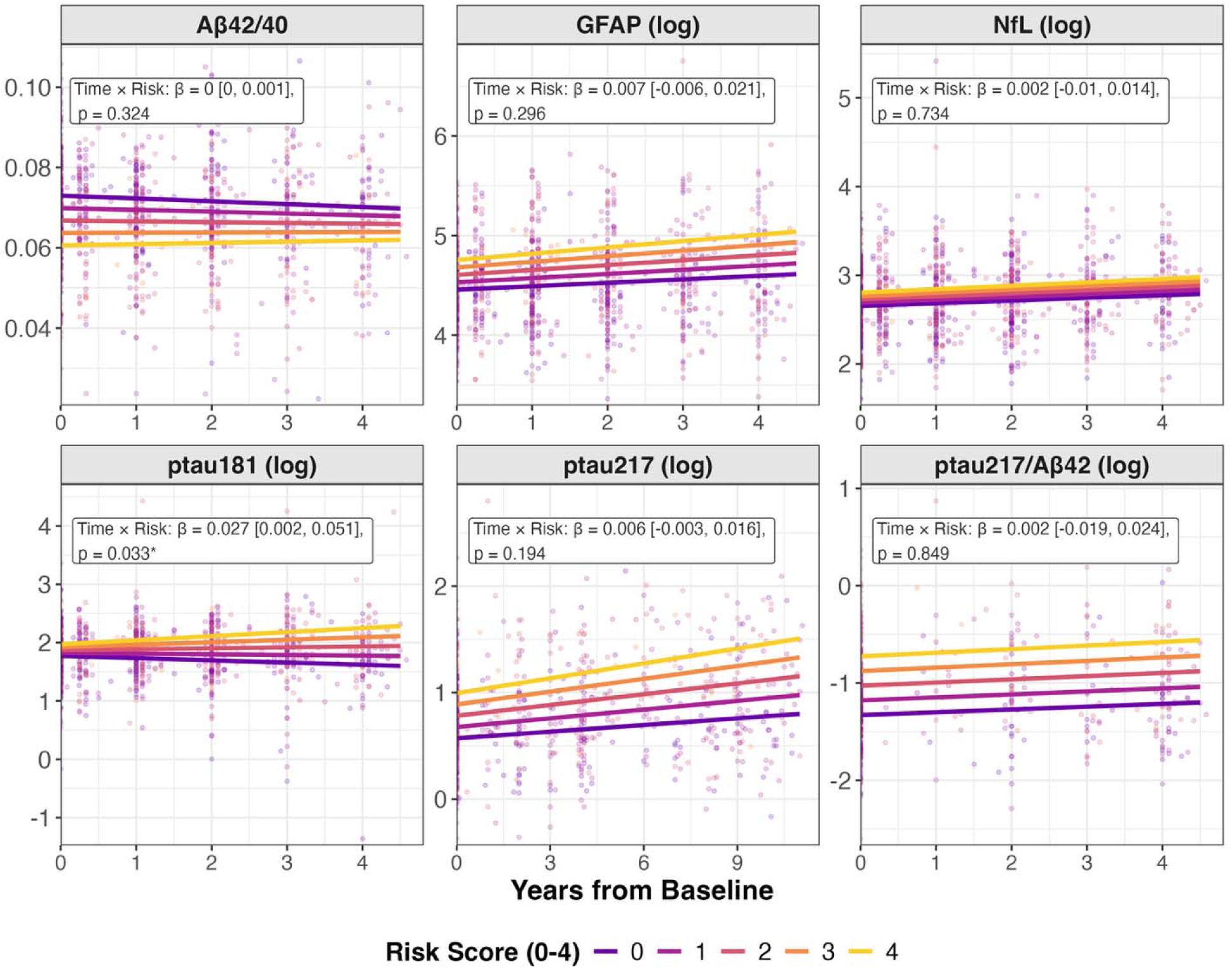
Predicted plasma biomarker trajectories by LIBRA2 composite AD risk score among PREVENT-AD participants. Points represent individual observations colored by risk score. Lines show population-average predicted trajectories from linear mixed-effects models (biomarker ∼ years × (composite risk score + age + sex + education) + (1 + years | participant)). Unadjusted p-values are shown; adjusted p-values can be found in Table S4. The composite risk score (0–4) reflects the count of elevated binary risk indicators: *APOE* ε4 carrier status, high PRS (top quartile), family history of AD (≥2 first-degree relatives), and high LIBRA2 score (1 SD above sample mean). Predictions are generated at a reference demographic profile (female sex, mean baseline age, mean years of education). Plasma biomarkers: Aβ_42/40_, GFAP, NfL, ptau_181_, ptau_217_, ptau_217_/Aβ_42_. Abbreviations: Aβ, amyloid beta; AD, Alzheimer’s disease; *APOE*, apolipoprotein E; β, beta coefficient; GFAP, glial fibrillary acidic protein; LIBRA2, Lifestyle for Brain Health Index; NfL, neurofilament light chain; p, p-value; PRS, polygenic risk score; ptau, phosphorylated tau; PREVENT-AD, Presymptomatic Evaluation of Experimental or Novel Treatments for Alzheimer’s Disease; SD, standard deviation.

## 4. Discussion

We examined the independent contributions of *APOE* ε4, PRS, family history burden, and polyexposure scores, and their cumulative effect as a composite score, on plasma biomarker levels and longitudinal trajectories in CU older adults. *APOE* ε4 emerged as the dominant predictor of plasma biomarker levels. Across cross-sectional and longitudinal analyses, higher composite risk burden was associated with elevated ptau_181_, ptau_217_, ptau_217_/Aβ_42_, and GFAP, lower Aβ_42/40_, and accelerated longitudinal accumulation of ptau_181_.

These findings are consistent with prior work on individual AD risk domains while extending them to a cumulative framework in a longitudinal cohort study of CU adults enriched for a family history of AD [16,17,28,29]. *APOE* ε4 showed the strongest and most consistent independent associations with elevated GFAP, ptau_181_, ptau_217_, and ptau_217_/Aβ_42_, and lower Aβ_42/40_ levels across cross-sectional and longitudinal multivariable frameworks, consistent with its well-established role in Aβ deposition and neuroinflammatory pathways [23,30]. Although nominal associations between PRS and GFAP and NfL were observed, no associations survived Hochberg correction. This may reflect the limited detectable neurodegeneration signal in a CU preclinical cohort and the moderate predictive power of the PRS [18].

Nominal CogDrisk associations with NfL, GFAP, and ptau_217_ levels suggest modifiable risk burden may influence neuroinflammatory/degenerative pathways and amyloid pathology prior to symptom onset [4,5,18,31,32]. However, these associations did not survive Hochberg correction and were not replicated in LIBRA2-based models, possibly reflecting the contribution of demographic factors in CogDrisk’s scoring, particularly age. The absence of an independent association between familial AD risk and plasma biomarker levels beyond genetic and lifestyle domains may reflect the enrichment of this cohort for AD family history, which may have restricted variability in this exposure and limited statistical power to detect independent familial effects.

The broader association of composite risk burden with plasma biomarker levels, despite weaker associations for several individual risk factors, suggests that cumulative risk may better capture subtle biological differences during the preclinical period than any single risk domain alone. Although composite risk burden was associated with biomarker levels at baseline and last visit, it was associated longitudinally only with ptau_181_ accumulation. Because ptau_217_ is generally more sensitive to amyloid pathology than ptau_181_, ptau_217_ may already have been elevated at baseline in this cohort, particularly given the inclusion of amyloid PET-positive participants, reducing its sensitivity to further risk-associated change during follow-up [4,31]. In contrast, the lower sensitivity of ptau181 to baseline amyloid burden may make it more responsive to longitudinal change. This interpretation is consistent with staging models in which ptau_217_ elevation precedes ptau_181_ [2,4,33]. Differences in assay sensitivity, dynamic range, follow-up duration, and biomarker assay frequency may also contribute to this pattern [34]. The absence of a longitudinal association with Aβ_42/40_ may reflect its comparatively limited peripheral sensitivity [35,36].

Several limitations warrant consideration when interpreting results. First, the small sample size may limit power to detect smaller effect sizes or non-additive interactions. Second, the potential interactions are not captured with an equal weighting composite model. Third, the PRS was constructed using only 74 genome-wide significant SNPs identified in Bellenguez et al. (2022), limiting the genetic variance captured and reducing power to detect associations with plasma biomarkers. Fourth, the analytical cohort is of European descent, limiting generalizability across ancestries where AD risk profiles and biomarker distributions may differ. Fifth, because PREVENT-AD is enriched for participants with a family history of AD, findings may not be generalizable to individuals without familial AD risk. Finally, though ptau_217_ measures were available over a longer observation window, its lower collection frequency reduced the number of longitudinal datapoints available for trajectory modeling, potentially limiting power to detect longitudinal changes.

Despite these limitations, our study has several strengths. First, PREVENT-AD’s deeply phenotyped longitudinal study design with annual biomarker collection enabled multivariable, cross-sectional, and longitudinal analyses within the same sample. Second, the use of two clinical risk scores with different risk item scoring reduces the likelihood that findings were an artifact of a singular measure of modifiable risk. Additionally, the simultaneous examination of genetic, familial, and modifiable lifestyle risk domains within a composite framework enabled assessment of independent contributions while accounting for intercorrelation. Finally, the use of a panel of plasma biomarkers that span amyloid, tau, and neuroinflammatory pathways allowed for interpretation of risk burden effects across multiple AD biological pathways.

A cumulative AD risk burden model– spanning genetic, familial, and modifiable clinical/lifestyle domains– supports the utility of multi-domain risk composites for preclinical AD risk stratification. External validation in diverse cohorts and examination of whether composite risk scores improve early identification of at-risk individuals that may progress to MCI status are important next steps. As plasma biomarkers become more clinically adopted, integrating composite risk profiles may help identify individuals most likely to benefit from emerging preventive interventions [16,17,37], particularly in diverse cohorts with more frequent longitudinal sampling.

## Supporting information

Supplementary Results

## Acknowledgments

We gratefully acknowledge the contributions of the PREVENT-AD participants, staff, and investigators to this study. PREVENT-AD was funded by the Alzheimer Society of Canada (NIG-17-08), the Alzheimer’s Association, and the Canadian Institutes of Health Research (CIHR; PJT-438655, PJT-367122, PJT-410106, PJT-463677).

## Data availability

Data used in preparation of this article were obtained from the Presymptomatic Evaluation of Experimental or Novel Treatments for Alzheimer’s Disease (PREVENT-AD) Registered repository available at https://registeredpreventad.loris.ca.

